# Rural-urban differences in gut microbiome among Vietnamese women

**DOI:** 10.64898/2026.10.04.26364707

**Authors:** Justin Guo, Huong T.T. Tran, Qiuyin Cai, Minh V. Hoang, Thuan V. Tran, Xiao-Ou Shu, Sang M. Nguyen

## Abstract

Gut microbiome plays an important role in human health. Urbanization has been suggested to alter the gut microbiome. We comprehensively evaluated differences in gut microbiome profiles of 76 urban and 81 rural female residents of Vietnam using shotgun metagenomic data. We evaluated rural-urban differences in gut microbial diversity, individual microbial taxa, and metabolic pathways. Rural residents had higher alpha and beta diversities in age-adjusted models (p<0.05). Additionally, significant rural-urban differences were observed for 35 microbial taxa in age-adjusted models (FDR<0.1). Rural residents had higher relative abundance of classes, orders, and families of *FGB2982* and *FGB38642* and strain *SGB15332* of the species *Faecalibacterium prausnitzii*, whereas urban residents had higher relative abundances of species *Bacteroides thetaiotaomicron, Coprobacter fastidiosus, Bilophila wadsworthia, Enterobacter cloacae, Enterobacter hormaechei, Klebsiella aerogenes*, *Dielma fastidiosa,* and *Holdemania massiliensis.* Notably, these associations were attenuated after further adjustment for socioeconomic characteristics, lifestyle, dietary factors, and comorbidity (FDR>0.1). In the fully adjusted model, significant rural-urban differences were observed in the abundance of 17 microbial taxa, including *Ruminococcus sp AF13 28* and *Prevotella pectinovora,* as well as in three metabolic pathways (FDR<0.1). This study suggests that urbanization-related factors may contribute to differences in the gut microbiome between urban and rural Vietnamese women.

## Introduction

The importance of the gut microbiome for human health is increasingly recognized [1]. A more diverse gut microbiome is typically more resilient to disturbances, increasing overall health [2]. In contrast, a microbiome with low biodiversity is positively associated with several chronic diseases [3–5]. Gut microbiome diversity can be affected by age, diet, physical activity, socioeconomic factors, well-being, and medication (especially antibiotics) [6–10]. The gut microbiota becomes compositionally unstable and less diverse with age-related declines in immunocompetence and with specific health conditions (e.g., inflammatory bowel disease, colorectal cancer, obesity, and extra-digestive diseases such as allergies) [11–13]. In addition, long-term geographic and lifestyle exposures, such as urban lifestyles, also shape gut microbial profiles [14]. Urban lifestyles are often associated with higher consumption of processed foods, lower fiber intake, greater antibiotic exposure, and more sedentary behavior, whereas rural populations tend to maintain traditional dietary patterns and an active lifestyle [15,16]. Urbanization was suggested as an important factor influencing population-level microbiota composition [17].

Vietnam has gone through significant economic change over the past four decades, accompanied by notable shifts in morbidity and mortality patterns. The country’s Gross Domestic Product (GDP) has grown rapidly: from USD 14.09 billion in 1985 to USD 514.7 billion in 2025 [18]. Alongside economic growth, the country has experienced rapid and widespread urbanization. Over the past 40 years, more and more Vietnamese have moved from rural areas to large urban centers. The rapid urbanization has been accompanied by lifestyle changes such as reduced physical activity and dietary shifts [19–22], which contributed to the rising prevalence of diabetes (from 1.5% in the 1990s to 4.1% in 2015) and overweight and obesity (from 2.3% in 1993 to 14.5% in 2015) among Vietnamese adults aged 25-64 [23–25]. It has also led to a significant shift in the health of Vietnamese citizens from communicable diseases to a high burden of non-communicable diseases (NCDs) [26].

The impact of urbanization (i.e., long-term geographic and urban lifestyle exposures) on the gut microbiota is thought to be considerable; however, few studies have evaluated differences in gut microbiome composition between urban and rural residents and their possible attributes in Vietnam. Recently, Hamdi & Truong et al. published the first report on how geography (urban vs. rural areas), diet, and aging shape the gut microbiota in 227 Vietnamese elderly [17]. They found that urban participants had higher richness and greater abundance of health-associated genera, including *Bifidobacterium*, *Fusicatenibacter*, and *Blautia*, while rural participants had a higher enrichment of beneficial butyrate-producing genera such as *Fusicatenibacter*, *Roseburia*, *Lachnospira,* and *Blautia*. However, this study was conducted on elderly Vietnamese aged ≥60 years and utilized data from 16S rRNA gene sequencing [17], In our previously reported study of 356 Vietnamese breast cancer patients using shotgun metagenomics, we found that income levels and fiber intake were significantly associated with the gut microbiome health index, alpha-diversity, and species belonging to the *Bifidobacterium*, *Prevotella*, and *Bacteroides* genera [27]. Studies of the general population with higher-resolution microbiome data are needed to better understand how urbanization influences the gut microbiome in Vietnam.

In the present study, we investigated differences in gut microbiome profiles between two groups of 164 Vietnamese women from urban and rural areas using shotgun metagenomic data. A better understanding of how urbanization affects gut microbiome diversity in Vietnam can inform public health and dietary guidelines tailored to Vietnam and other low- and middle-income countries with similar economic transitions.

## Materials and Methods

### Study Population

We analyzed shotgun metagenomic sequencing data from 164 women enrolled in a cross-sectional study of 1,035 participants, recruited between January and September 2018 in Hanoi (urban area) and Hai Duong province (rural area), Vietnam.

Briefly, a multistage cluster sampling method was applied to select participants aged 40- 9 years in one urban and one rural province of Vietnam (Hanoi and Hai Duong, respectively). First, local health staff obtained a list of families that had at least two people (one man and one woman) aged 40-79 years in each area. A sample of 530 households, including 250 in Hanoi and 280 in Hai Duong province, was selected randomly. Participants were recruited into the study if they met the following inclusion criteria: (1) currently living in the selected areas and present during the recruitment times; (2) aged 40 to 79 years at the time of the survey; and (3) were able to provide informed consent. We excluded participants if they (1) received any treatment at a health facility or were diagnosed with severe diseases (e.g., cancer, myocardial infarction, stroke, etc.) less than 12 months ago; (2) were determined to be physically or mentally incapable of completing the interview by research staff; and (3) had physical deformities or deafness.

Interviewers, trained by the Vietnam National Cancer Institute and the Hanoi University of Public Health, interviewed participants and directly contacted potential eligible participants in their households to introduce the study and invited one man and one woman who met the above-mentioned eligibility criteria to participate. In-person interviews began only after obtaining the patient’s consent form, using a structured questionnaire administered via the Research Electronic Data Capture (REDCap) mobile application [28]. The response rate in the current study was approximately 98.0% in both areas, resulting in a total of 1,035 participants, equally divided into urban and rural areas. The study was approved by the Institutional Review Boards of the participating institutions, including Hanoi University of Public Health and Vietnam National Cancer Institute.

Residence in urban and rural areas was classified based on where the participant had lived for most of the prior 10 years at the time of enrollment. Information on participants’ demographic characteristics (age, education, household income, family size), menopausal status, body mass index (BMI), physical activity, sitting time, sleep duration, clinical history, oral health, medication use, dietary frequency, and medication and supplement use was collected at study enrollment.

### Stool sample collection, Microbiome DNA extraction, and metagenomic sequencing

Stool samples were collected from 490 women and 395 men using fecal occult blood test (FOBT) cards at study enrollment, following a standard protocol. Two FOBT cards were provided to each participant, along with clear instructions for collecting stool samples at home. The stool samples were transferred to the Vietnam National Cancer Institute research laboratory and then stored in a -80 °C freezer.

In the current study, stool samples from 164 women were randomly selected by residence area and 5-year age groups, and DNA was extracted using the DNeasy PowerSoil Kit (Qiagen, Carlsbad, CA, USA) following the manufacturer’s protocols. The DNBSEQ Short-read library preparation kit was used to build sequencing libraries from DNA samples for shotgun metagenomic sequencing. Sequencing was conducted in 150 bp paired-end mode on the MGISEQ-2000 at BGI Americas (Cambridge, MA, USA). DNA extraction, library preparation, and sequencing of all samples were performed in one batch.

Sequencing reads were processed with Trimmomatic v0.39 to trim low-quality bases, after which reads shorter than 105 nucleotides (70% of the original read length) were discarded [29]. Then, Bowtie2 v2.3.0 was used to remove reads that could be mapped to the human genome (GRCh38) [30]. Clean reads were subjected to taxonomic profiling using MetaPhlAn 4.1.1 [31]. HUMAnN 3.6 was utilized to perform functional profiling of the gut microbiome using clean reads and the UniRef90 comprehensive protein database as a reference. We estimated the relative abundance of gut microbial metabolic pathways [32].

The current study included 157 female participants in the final analysis after excluding 7 participants who reported antibiotic use within 24 hours before sample collection, given the impact of antibiotic therapy on the gut microbiome.

### Statistical Analysis

Descriptive statistics for patients’ demographic characteristics were computed as percentages for categorical variables and as means and standard deviations (SDs) for continuous variables. Urban vs rural differences in participant characteristics were compared using Wilcoxon rank-sum tests for continuous variables and Pearson chi-square or Fisher exact tests for categorical variables, as appropriate.

We measured gut microbiome overall microbial richness (α-diversity) using the observed richness index, Shannon index, inverse Simpson index, and Faith’s phylogenetic diversity (PD) index, and similarities or dissimilarities between communities (i.e., samples) by β-diversity using the Bray-Curtis dissimilarity matrix, weighted UniFrac distance matrix, and unweighted UniFrac distance, using the R functions *VEGAN::DIVERSITY* and *VEGAN::VEGDIST*, respectively.[33]

Rural-urban differences in gut microbiome α-diversity indexes were evaluated via multivariable linear regression models. The mean difference (β coefficients) and standard error (SE) were estimated and presented in three sets of analyses. Model 1 adjusted for age only; Model 2 additionally adjusted for education (under high school vs. High school or higher), income (< VND 100 million vs. > VND 100 million), regular exercise (yes/no), total physical activity (continuous - MET-h/week), calcium/vitamin supplement use (yes/no), and frequency of consuming beef (<2 times/week vs. ≥2 times/week), pork (≤7 times/week vs. >7 times/week), rice (≤14 times/week vs. >14 times/week), and vegetables (<14 times/week vs. ≥14 times/week. Model 3 further adjusted for comorbidity (i.e., any diagnoses of diabetes, high blood pressure, hyperlipidemia, coronary heart disease, stroke, chronic gastritis, ulcer, chronic pulmonary disease, and arthritis), number of teeth lost due to health reasons, and receipt of antibiotics (oral or injectable) during the past 3 months (yes/no). The permutational Multivariate Analysis of Variance (PERMANOVA) test was used to assess differences in β-diversity between rural and urban residents [34]. R-squared values from PERMANOVA tests for residence were derived from models adjusted for the aforementioned covariates, with 999 permutations, using the R function *vegan* [33]. All statistical analyses for α- and β-diversities were performed using two-sided tests, and associations with *P*<0.05 were considered statistically significant.

Rural-urban differences in gut microbial taxa abundance and metabolic pathways were assessed using differential abundance analysis (LinDA, R package “*MicrobiomeStat*” version 1.5.0 [35]), which was adjusted for the potential confounders as mentioned above. Log_2_ Fold Change (Log_2_ FC), standard error (SE), and p-values for individual taxa/pathways were produced. We limited our analysis to gut microbial taxa and metabolic pathways with prevalence >10% and median relative abundance >0.001%. In this study, common taxa were defined as present in >50% of samples; rare taxa were defined as present in <50% of samples. The false discovery rate (FDR) was calculated at each taxonomic level using common and rare taxa to control for multiple testing. An association with an FDR-corrected p-value <0.1 was considered statistically significant. All statistical analyses were conducted in R version 4.5.2.

## Results

### Participant characteristics

Among 157 female participants, 76 (48.4%) were urban residents and 81 (51.6%) were rural residents. The mean age (SD) was 52.9 (7.5) years in urban residents and 52.4 (8.7) years in rural residents. Compared with rural residents, urban residents were more likely to have higher educational attainment, higher household income, and larger family sizes. No differences were observed between the two groups in marital status or menopausal status. However, regular leisure-time exercise was more common among urban residents (72.4% vs 28.4%; p<0.001), whereas total physical activity was higher among rural residents (197.5 ± 109.3 vs 113.9 ± 70.0 MET-hours/week; p<0.001). BMI, BMI levels, sitting time, and sleep duration were similar between the two groups (**Table 1 & Supplementary Table 1)**.

**Table 1.** Participant characteristics, lifestyle, disease history, medication use, and dietary variables by residence (N=157)

| Characteristic | Overall<br>N = 157 | Urban<br>N = 76 | Rural<br>N = 81 | P<br>value |
| --- | --- | --- | --- | --- |
| **Demographics and socioeconomic factors** |  |  |  |  |
| Age, years | 52.6 ± 8.1 | 52.9 ± 7.5 | 52.4 ± 8.7 | 0.508 |
| Age ≥60 years | 31 (19.7%) | 14 (18.4%) | 17 (21.0%) | 0.795 |
| Married | 154 (98.1%) | 74 (97.4%) | 80 (98.8%) | 0.611 |
| College or higher education | 28 (17.8%) | 20 (26.3%) | 8 (9.9%) | 0.012 |
| Household income >150 million VND | 21 (13.4%) | 14 (18.4%) | 7 (8.6%) | 0.038 |
| **Lifestyle and anthropometrics** |  |  |  |  |
| Postmenopausal | 88 (56.1%) | 44 (57.9%) | 44 (54.3%) | 0.652 |
| BMI, kg/m<sup>2</sup> | 22.3 ± 2.7 | 22.2 ± 2.5 | 22.3 ± 2.9 | 0.993 |
| Regular exercise | 78 (49.7%) | 55 (72.4%) | 23 (28.4%) | <0.001 |
| Total physical activity, MET-hours/week | 157.0 ± 101.2 | 113.9 ± 70.0 | 197.5 ± 109.3 | <0.001 |
| Sitting, hours/day | 3.4 ± 2.7 | 3.2 ± 2.9 | 3.5 ± 2.6 | 0.453 |
| Sleep, hours/day | 6.9 ± 1.8 | 6.8 ± 1.8 | 7.0 ± 1.7 | 0.216 |
| **Long-term medication use** |  |  |  |  |
| Multiple vitamins | 24 (15.3%) | 17 (22.4%) | 7 (8.6%) | 0.017 |
| Calcium and/or vitamin D | 40 (25.5%) | 26 (34.2%) | 14 (17.3%) | 0.015 |
| Any antibiotics (oral or injectable) during past 3 months | 29 (18.5%) | 10 (13.2%) | 19 (23.5%) | 0.097 |
| **Dietary frequency** |  |  |  |  |
| Beef ≥2 times/week | 34 (21.7%) | 24 (31.6%) | 10 (12.3%) | <0.001 |
| Pork >7 times/week | 48 (30.6%) | 14 (18.4%) | 34 (42.0%) | 0.002 |
| Rice/sticky rice >14 times/week | 38 (24.2%) | 9 (11.8%) | 29 (35.8%) | <0.001 |
| Bread ≥2 times/week | 53 (33.8%) | 31 (40.8%) | 22 (27.2%) | <0.001 |
| Vegetables ≥14 times/week | 56 (35.7%) | 14 (18.4%) | 42 (51.9%) | <0.001 |
| Nuts/peanuts >2 times/week | 42 (26.8%) | 23 (30.3%) | 19 (23.5%) | 0.002 |
| Fruits ≥7 times/week | 47 (29.9%) | 27 (35.5%) | 20 (24.7%) | 0.027 |
| Deep fried/grilled foods ≥2 times/week | 46 (29.3%) | 34 (44.7%) | 12 (14.8%) | <0.001 |
| **History of disease and oral health** |  |  |  |  |
| Diabetes | 9 (5.7%) | 3 (3.9%) | 6 (7.4%) | 0.497 |
| High blood pressure | 27 (17.2%) | 11 (14.5%) | 16 (19.8%) | 0.381 |
| Hyperlipidemia | 39 (24.8%) | 23 (30.3%) | 16 (19.8%) | 0.128 |
| Coronary heart disease | 12 (7.6%) | 5 (6.6%) | 7 (8.6%) | 0.627 |
| Chronic gastric/Ulcer | 55 (35.0%) | 31 (40.8%) | 24 (29.6%) | 0.143 |
| Chronic pulmonary disease | 17 (10.8%) | 11 (14.5%) | 6 (7.4%) | 0.154 |
| Arthritis | 55 (35.0%) | 35 (46.1%) | 20 (24.7%) | 0.005 |
| Any tooth loss | 71 (45.2%) | 25 (32.9%) | 46 (56.8%) | 0.003 |
| Number of teeth lost due to health reasons | 1.6 ± 2.8 | 1.2 ± 2.9 | 1.9 ± 2.7 | 0.019 |

<sup>1</sup>n (%); Mean±SD<sup>2</sup>Fisher's exact test; Pearson's Chi-squared test; Wilcoxon rank sum test

Several self-reported medical conditions were more frequent among urban residents, including ulcers (25.0% vs. 9.9%; p=0.012) and arthritis (46.1% vs. 24.7%; p=0.005). Conversely, tooth loss was more common among rural participants (56.8% vs 32.9%; p=0.003) compared with urban residents. No differences were observed between the two groups in diabetes, high blood pressure, hyperlipidemia, coronary heart disease/stroke, chronic gastritis, and chronic pulmonary disease (**Table 1 & Supplementary Table 2).** Use of multiple vitamins (24.4% vs. 8.6%; p=0.017) and calcium or vitamin D (34.2% vs. 17.3%; p=0.015) was more common among urban residents than among rural residents. Antibiotic use in the prior 3 months was more common in rural participants, but the difference was not statistically significant (**Table 1 & Supplementary Table 3)**

High-frequency consumption of beef, bread, fruit, nuts or peanuts, and deep-fried or grilled food was observed among urban residents, whereas higher-frequency intake of pork, rice or sticky rice, and vegetables was observed among rural residents (All p <0.05; **Table 1 & Supplementary Table 4**).

### Rural-urban differences in gut microbial α- and β-diversity

In age-adjusted models, rural residence was significantly associated with higher observed richness (β[se] = 24.26 [11.18]; p = 0.031) and higher Faith’s phylogenetic diversity (β[se] = 2.35 [1.12]; p = 0.037). These associations were attenuated after accounting for demographic and lifestyle covariates and remained nonsignificant in the fully adjusted model. Shannon diversity, Pielou evenness, and inverse Simpson diversity did not differ significantly by residence in any model (**Table 2**).

**Table 2.**
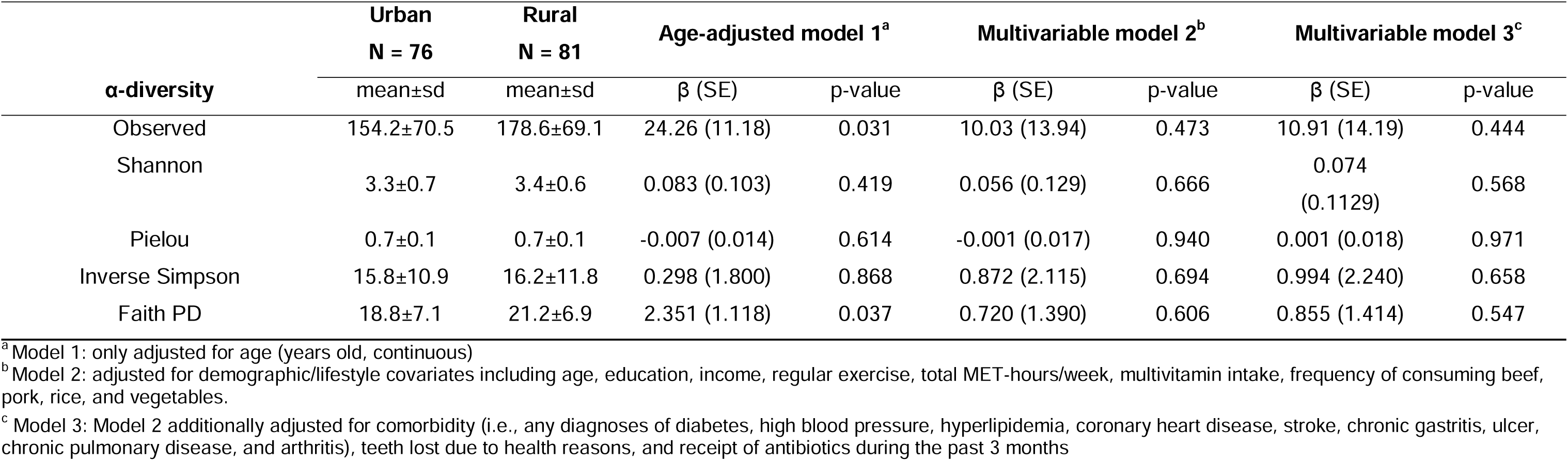
Rural-urban differences in gut microbiome α-diversity (N=157)

|  | **Urban** | **Rural** | **Age-adjusted model 1<sup>a</sup>** |  | **Multivariable model 2<sup>b</sup>** |  | **Multivariable model 3<sup>c</sup>** |  |
| --- | --- | --- | --- | --- | --- | --- | --- | --- |
|  | **N = 76** | **N = 81** |  |  |  |  |  |  |
| **α-diversity** | mean±sd | mean±sd | $\beta$ (SE) | p-value | $\beta$ (SE) | p-value | $\beta$ (SE) | p-value |
| Observed | 154.2±70.5 | 178.6±69.1 | 24.26 (11.18) | 0.031 | 10.03 (13.94) | 0.473 | 10.91 (14.19) | 0.444 |
| Shannon | 3.3±0.7 | 3.4±0.6 | 0.083 (0.103) | 0.419 | 0.056 (0.129) | 0.666 | 0.074<br>(0.1129) | 0.568 |
| Pielou | 0.7±0.1 | 0.7±0.1 | -0.007 (0.014) | 0.614 | -0.001 (0.017) | 0.940 | 0.001 (0.018) | 0.971 |
| Inverse Simpson | 15.8±10.9 | 16.2±11.8 | 0.298 (1.800) | 0.868 | 0.872 (2.115) | 0.694 | 0.994 (2.240) | 0.658 |
| Faith PD | 18.8±7.1 | 21.2±6.9 | 2.351 (1.118) | 0.037 | 0.720 (1.390) | 0.606 | 0.855 (1.414) | 0.547 |

<sup>a</sup> Model 1: only adjusted for age (years old, continuous)
<sup>b</sup> Model 2: adjusted for demographic/lifestyle covariates including age, education, income, regular exercise, total MET-hours/week, multivitamin intake, frequency of consuming beef, pork, rice, and vegetables.
<sup>c</sup> Model 3: Model 2 additionally adjusted for comorbidity (i.e., any diagnoses of diabetes, high blood pressure, hyperlipidemia, coronary heart disease, stroke, chronic gastritis, ulcer, chronic pulmonary disease, and arthritis), teeth lost due to health reasons, and receipt of antibiotics during the past 3 months

Rural-Urban difference explained a small but statistically significant proportion of the between-sample variation in age-adjusted beta diversity for Bray-Curtis dissimilarity (R² = 1.5%; p = 0.003) and unweighted UniFrac distance (R² = 1.3%; p = 0.025) but not for weighted UniFrac distance. After additional adjustment for demographic, lifestyle, and comorbid conditions, associations with all three beta-diversity metrics lost their statistical significance (**Table 3**)

**Table 3.**
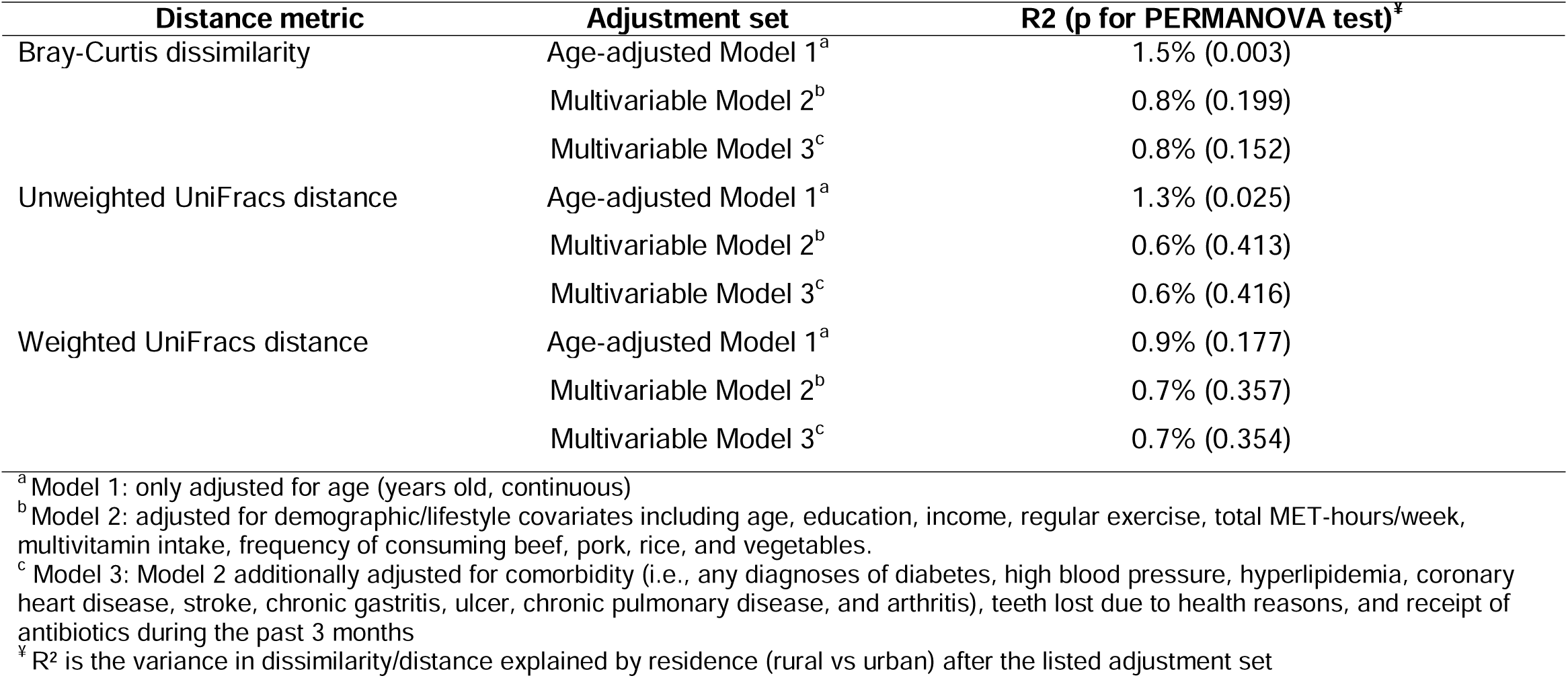
PERMANOVA test of differences in beta diversity by residence (N=157)

| Distance metric | Adjustment set | R2 (p for PERMANOVA test)<sup>‡</sup> |
| --- | --- | --- |
| Bray-Curtis dissimilarity | Age-adjusted Model 1<sup>a</sup> | 1.5% (0.003) |
|  | Multivariable Model 2<sup>b</sup> | 0.8% (0.199) |
|  | Multivariable Model 3<sup>c</sup> | 0.8% (0.152) |
| Unweighted UniFrac distance | Age-adjusted Model 1<sup>a</sup> | 1.3% (0.025) |
|  | Multivariable Model 2<sup>b</sup> | 0.6% (0.413) |
|  | Multivariable Model 3<sup>c</sup> | 0.6% (0.416) |
| Weighted UniFrac distance | Age-adjusted Model 1<sup>a</sup> | 0.9% (0.177) |
|  | Multivariable Model 2<sup>b</sup> | 0.7% (0.357) |
|  | Multivariable Model 3<sup>c</sup> | 0.7% (0.354) |

<sup>a</sup> Model 1: only adjusted for age (years old, continuous)
<sup>b</sup> Model 2: adjusted for demographic/lifestyle covariates including age, education, income, regular exercise, total MET-hours/week, multivitamin intake, frequency of consuming beef, pork, rice, and vegetables.
<sup>c</sup> Model 3: Model 2 additionally adjusted for comorbidity (i.e., any diagnoses of diabetes, high blood pressure, hyperlipidemia, coronary heart disease, stroke, chronic gastritis, ulcer, chronic pulmonary disease, and arthritis), teeth lost due to health reasons, and receipt of antibiotics during the past 3 months
<sup>‡</sup> R<sup>2</sup> is the variance in dissimilarity/distance explained by residence (rural vs urban) after the listed adjustment set

### Rural-Urban differences in gut microbial composition

Among the 157 study stool samples, 16 phyla, 329 classes, 356 orders, 416 families, 1,234 genera, 2,204 species, and 2,352 strains were identified. At the phylum level, *Bacteroidota, Firmicutes, Proteobacteria,* and *Actinobacteria* had the highest relative abundances (**Figure 1**). After excluding rare taxa with a prevalence of <10% in all samples, a total of 2,869 gut microbial taxa, including 9 phyla, 57 classes, 64 orders, 85 families, 203 genera, 364 species, and 393 strains, were included in further evaluation.

**Figure 1.**
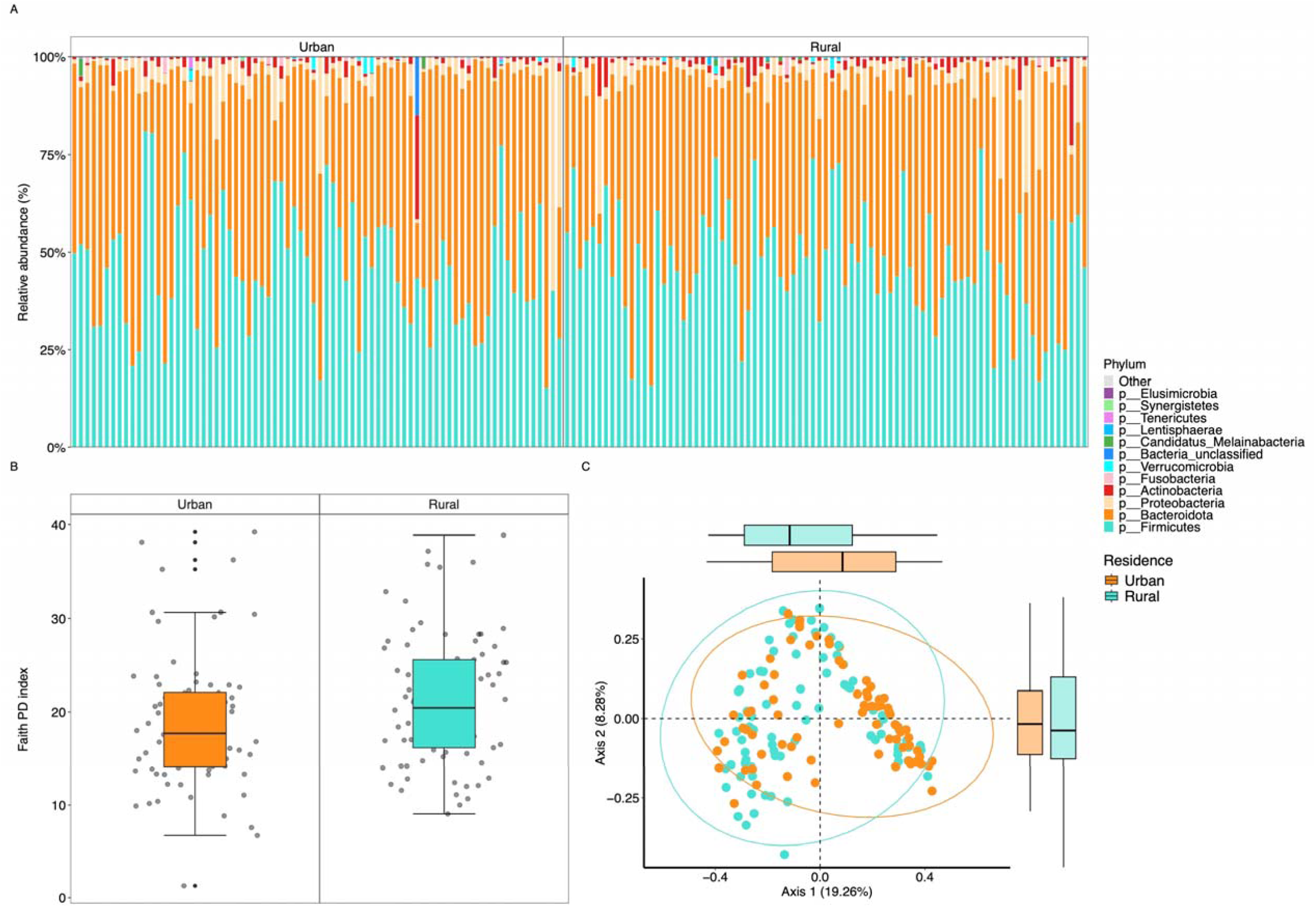
(A) Relative abundance at phylum levels by residence. (B) Alpha diversity measured in terms of Faith’s phylogenetic diversity (FD) index) by residence. (C) PCoA of Bray-Curtis diversity by residence.

In age-adjusted models, 35 microbial taxa differed significantly by residence, including 8 with higher abundance in rural residents and 27 with higher abundance in urban residents (FDR <0.1). Rural-enriched microbial taxa included classes, orders, and families of *FGB2982* and *FGB38642,* as well as class *CFGB1474* and the strain *SGB15332* of the species *Faecalibacterium prausnitzii*, all of which belong to the phylum Firmicutes. Urban-enriched microbial taxa included four taxa belonging to phylum *Bacteroidota* (i.e., species *Bacteroides thetaiotaomicron* and it strain *SGB1861,* species *Coprobacter fastidiosus* and its strain *SGB1963),* eight taxa belonging to phylum *Proteobacteria* (i.e., species *Bilophila wadsworthia* and its strain *SGB15452,* species *Enterobacter cloacae* and its strain *SGB10131,* species *Enterobacter hormaechei* and its strain *SGB10130,* and species *Klebsiella aerogenes* and its strain *SGB10116*), and 15 taxa belonging to phylum *Firmicutes* (eg. taxa-related class *CFGB1292,* species *Dielma fastidiosa, Holdemania massiliensis* and their strains, and strains of *Blautia hansenii, Enterocloster bolteae,* and *Wansuia hejianensis*). After additional adjustment for socioeconomic characteristics, lifestyle, dietary factors, and comorbidity, all associations lost their statistical significance (**Figure 2 & Supplementary Table 5**).

**Figure 2.**
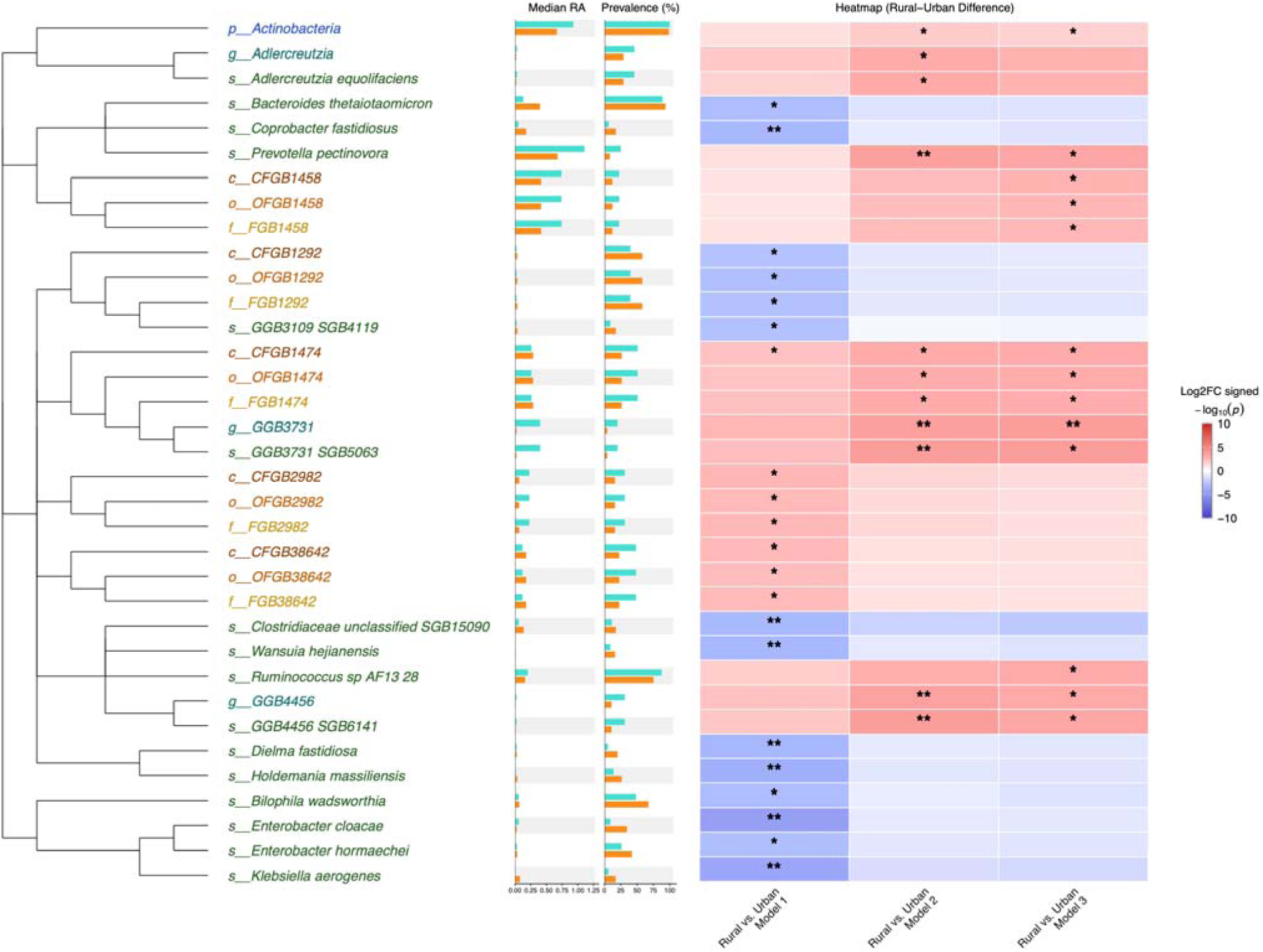
Rural-Urban differences in gut microbial composition (N=157). Model 1: adjusted for age; Model 2: adjusted for demographic/lifestyle covariates, including age, education, income, regular exercise, total MET-hours/week, calcium/multivitamin intake, and the frequency of beef, pork, rice, and vegetable consumption. Model 3: Model 2 additionally adjusted for comorbidity (i.e., any diagnoses of diabetes, high blood pressure, hyperlipidemia, coronary heart disease, stroke, chronic gastritis, ulcer, chronic pulmonary disease, and arthritis), teeth lost due to health reasons, and receipt of antibiotics during the past 3 months. In bar charts by residence, turquoise indicates rural and tangerine indicates urban. Significant heatmap cells are indicated by FDR-corrected q-values (* 0.05≤FDR q <0.10, ** <0.05), and association directions are shown by color, with red indicating positive log2FC and blue indicating negative log2FC. Abbreviations: FC: Fold change; FDR: False discovery rate; RA = relative abundance.

In fully adjusted models (Model 3), 17 microbial taxa showed significant differences between rural and urban areas (FDR < 0.1). Significantly higher enrichment of the species *Prevotella pectinovora* (Log2 FC [SE] = 1.71 [0.48]; p=0.001; FDR=0.061) and its strain *SGB1662* (Log2 FC [SE] = 1.71 [0.48]; p=4.92×10^-4^; FDR=0.067) was found among rural residents. Additionally, rural residents had a significantly higher enrichment of the class *CFGB1458* (Log2 FC [SE] = 2.39 [0.76]; p=0.002; FDR=0.056), and order *OFGB1458* (Log2 FC [SE] = 2.37 [0.76]; p=0.002; FDR=0.074), which were driven by their family *FGB1458* (Log2 FC [SE] = 2.34 [0.76]; p=0.003; FDR=0.093). Rural residents had a significantly higher abundance of the class *CFGB1474* (Log2 FC [SE] = 2.56 [0.77]; p=0.001; FDR=0.056), order *OFGB1474* (Log2 FC [SE] = 2.54 [0.77]; p=0.001; FDR=0.074), and family *FGB1474* (Log2 FC [SE] = 2.51 [0.76]; p=0.001; FDR=0.089), which was driven by their genus *GGB3731* (Log2 FC [SE] = 1.79 [0.47]; p=1.83×10^-4^; FDR=0.029), species *GGB3731_SGB5063* (Log2 FC [SE] = 1.81 [0.47]; p=1.65×10^-4^; FDR=0.051), and strain *SGB5063* (Log2 FC [SE] = 1.81 [0.47]; p=1.56×10^-4^; FDR=0.054). Moreover, the abundance of the genus *GGB4456* (Log2 FC [SE] = 1.77 [0.51]; p=0.001; FDR=0.061) and species *GGB4456_SGB6141* (Log2 FC [SE] = 1.79 [0.51]; p=0.001; FDR=0.061), its strain *SGB6141* (Log2 FC [SE] = 1.79 [0.51]; p=0.001; FDR=0.067) was significantly higher among rural residents. The abundance of the species *Ruminococcus_sp_AF13_28* (Log2 FC [SE] = 3.02 [0.89]; p=0.001; FDR=0.076) and its strain *SGB4834* (Log2 FC [SE] = 3.03 [0.89]; p=0.001; FDR=0.073) was significantly higher among rural residents. Finally, rural residents had a significantly higher abundance of the phylum *Actinobacteria* (Log2 FC [SE] = 0.86 [0.37]; p=0.023; FDR=0.091), driven by the genus *Adlercreutzia*, the species *Adlercreutzia equolifaciens*, and the strain *SGB14798*. However, these associations with the three taxa were only significant at FDR<0.1 in Model 2 when adjusted for socioeconomic characteristics, lifestyle, and dietary factors, but they lost statistical significance in the fully adjusted Model 3, with additional adjustments for comorbidity, oral health, and antibiotic use (**Figure 2 & Supplementary Table 5**).

### Rural-Urban differences in gut microbial metabolic pathways

A total of 635 gut microbial metabolic pathways were identified, and their relative abundances were estimated. After excluding microbial metabolic pathways that were present in <10% of samples, a total of 421 gut microbial metabolic pathways were included in the analysis. We only found three microbial metabolic pathways associated with residence (p < 0.05, and FDR < 0.1) in the fully adjusted model (Model 3): rural residents had a significantly higher abundance of L-glutamate degradation VIII to propanoate (PWY-5088: Log2 FC [SE] = 1.12 [0.32]; p=0.001; FDR=0.033) and a significantly lower abundance of sucrose degradation II (PWY-3801: Log2 FC [SE] = -0.78 [0.27]; p=0.005; FDR=0.084) and superpathway of anaerobic sucrose degradation (PWY-7345: Log2 FC [SE] = -0.79 [0.27]; p=0.005; FDR=0.084). We also observed 28 microbial metabolic pathways associated with residence (p<0.05) across three models, but none reached statistical significance at FDR<0.1 (**Figure 3 and Supplementary Table 6**).

**Figure 3.**
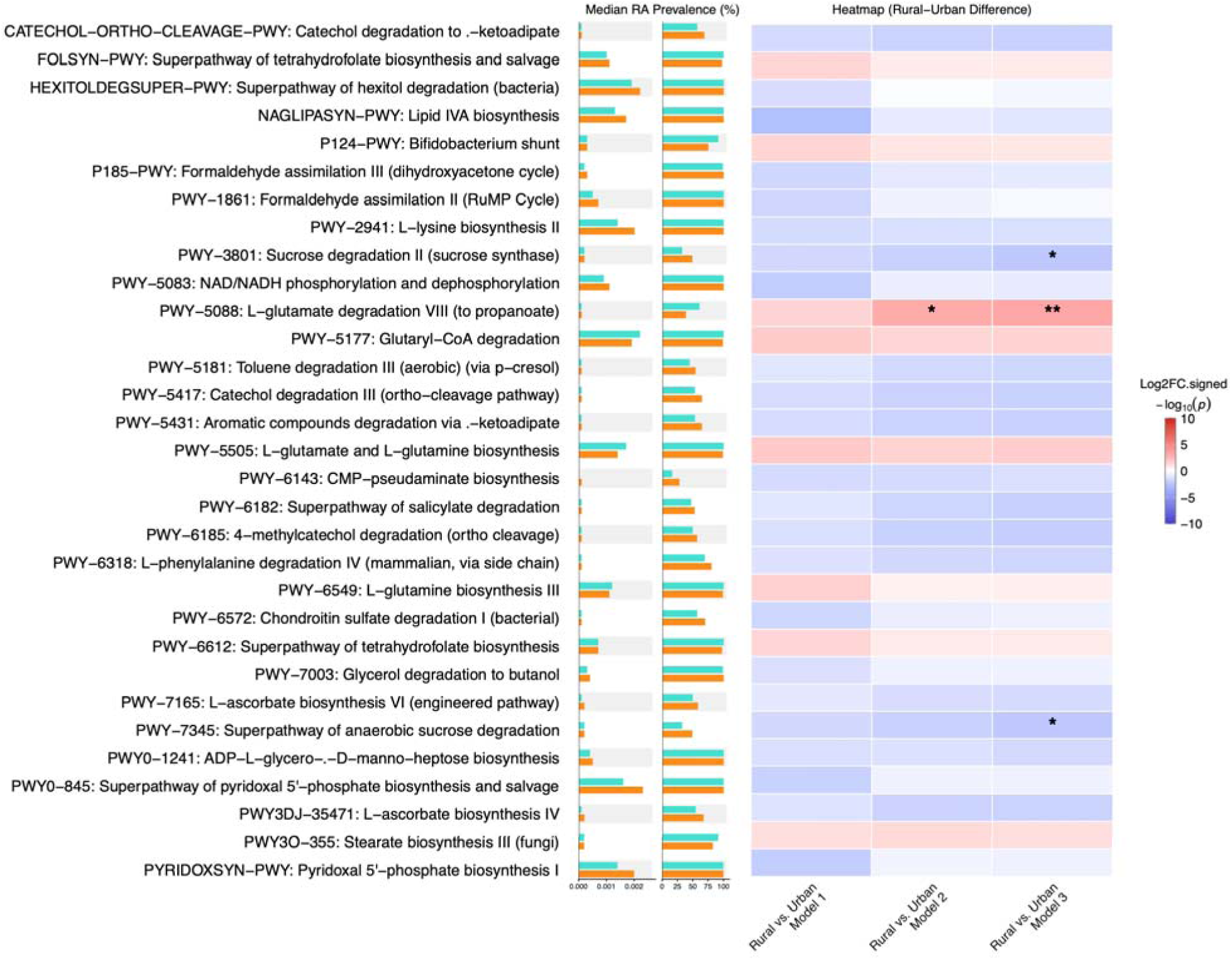
Rural-urban differences in gut microbial metabolic pathways (N=157). Model 1: adjusted for age; Model 2: adjusted for demographic/lifestyle covariates, including age, education, income, regular exercise, total MET-hours/week, calcium/multivitamin intake, and the frequency of beef, pork, rice, and vegetable consumption. Model 3: Model 2 additionally adjusted for comorbidity (i.e., any diagnoses of diabetes, high blood pressure, hyperlipidemia, coronary heart disease, stroke, chronic gastritis, ulcer, chronic pulmonary disease, and arthritis), teeth lost due to health reasons, and receipt of antibiotics during the past 3 months. In bar charts by residence, turquoise indicates rural and tangerine indicates urban. Significant heatmap cells are indicated by FDR-corrected q-values (* 0.05≤FDR q <0.10, ** <0.05), and association directions are shown by color, with red indicating positive log2FC and blue indicating negative log2FC. Abbreviations: FC: Fold change; FDR: False discovery rate; RA = relative abundance.

## Discussion

In this community-based study of Vietnamese women, we observed that rural residents had higher alpha diversity, including microbial richness and phylogenetic diversity, beta diversity, and enrichment of several microbial taxa belonging to the phylum *Firmicutes* (i.e., classes, orders, and families of *CFGB2982, CFGB38642,* and strain *SGB15332* of the species *Faecalibacterium prausnitzii).* Additionally, we also observed that urban residents had enrichment of microbial taxa belonging to phylum *Bacteroidota, Proteobacteria,* and *Firmicutes* (e.g., species *Bacteroides thetaiotaomicron, Coprobacter fastidiosus, Bilophila wadsworthia, Enterobacter cloacae, Enterobacter hormaechei, Klebsiella aerogenes*, *Dielma fastidiosa, Holdemania massiliensis,* and strains of *Blautia hansenii, Enterocloster bolteae,* and *Wansuia hejianensis*). However, most of these associations were attenuated after adjustment for socioeconomic characteristics, lifestyle, dietary factors, and comorbidity with the exception of observed enrichment in species *Prevotella pectinovora*, strain *SGB1662*, species *Ruminococcus_sp_AF13_28,* strain *SGB1662,* class *CFGB1474*, order *OFGB1474*, and family *FGB1474,* genus *GGB3731,* species *GGB3731_SGB5063,* strain SGB*5063,* genus *GGB4456,* species *GGB4456_SGB6141,* strain *SGB6141,* class *CFGB1458,* order *OFGB1458,* family *FGB1458,* and phylum *Actinobacteria* among rural participants. This study suggested that changes accompanying urbanization may be involved in shaping the gut microbiome profile of Vietnamese women and underscores the complex interplay of lifestyle, socioeconomic, and dietary factors, as well as comorbidity, in gut microbial communities.

Urbanization and geography of residency may influence gut microbiota composition [17]. Pronounced differences in bacterial species assemblages and functional gene repertoires were observed among inhabitants of a USA metropolitan area and residents of rural communities in Malawi and Venezuela [36]. A study of the fecal microbiota of 314 healthy young adults across 20 rural and urban cohorts from nine provinces of China reported significant structural segregation of the gut microbiota, clustering mainly by geography and ethnicity and less so by lifestyle [36]. Another study of 7,009 individuals across 14 districts in one province of China, using 16S rRNA sequencing, suggests that urban populations have reduced microbial diversity and an increased prevalence of virulence and antibiotic-resistance genes [37]. Similarly, studies of African individuals have reported that rural populations consistently exhibit greater diversity and stability in their gut microbiota than urban populations [38,39]. The geographic differences in microbial diversity and composition have been consistently highlighted in studies in India, Kazakhstan, Malaysia, and Thailand [40–44].

In line with these studies, our study found that rural residents had higher microbial richness and phylogenetic diversity, along with enrichment of the strain *SGB15332* of the species *Faecalibacterium prausnitzii. F. prausnitzii* is known as a beneficial gut bacterium that primarily produces butyrate, a short-chain fatty acid (SCFA), and bioactive anti-inflammatory molecules, and is recognized as a key player in the gut health and physiology of the host [45,46]. Vietnamese people typically eat rice and plant-based diets (i.e., high-fiber intake), moderate amounts of animal protein, and fermented foods rich in natural probiotics, with these dietary habits more prevalent in rural areas [47]. In our current study, women living in rural areas reported consuming vegetables and rice at least twice per day more frequently, which may explain their greater microbial richness. Although women living in urban areas consumed vegetables less frequently than rural residents, they consumed fruits, nuts/peanuts, and yogurt more frequently, which may explain their enrichment of microbial taxa belonging to the phyla *Bacteroidota* and *Firmicutes* (e.g., species such as *Bacteroides thetaiotaomicron, Coprobacter fastidiosus,* and strains of *Blautia hansenii*)*. Bacteroides thetaiotaomicron* has been shown to positively affect anti-inflammatory immune responses. Several studies also report microbial dysregulation in patients with Inflammatory Bowel Disease (IBD), along with reductions in the abundance of *Bacteroides* species, including *Bacteroides thetaiotaomicron* [48,49]. Similarly, *Coprobacter fastidiosus* is associated with a lower risk of Inflammatory Bowel Disease (IBD) and ulcerative colitis (UC) [50,51]. *Blautia hansenii, a Blautia species,* produces acetic acid and butyric acid, and was significantly and negatively associated with visceral fat area. *Blautia* is reportedly less prominent in obese children, diabetic adults, and pediatric patients, as well as subjects with other diseases, such as rectal cancer and rheumatoid arthritis, than in healthy subjects [52]. Similar enrichment of *Bacteroides* and *Blautia* has also been observed in Japanese and Korean cohorts residing in highly urbanized areas [53–55].

Urban residents had an enrichment of microbial taxa belonging to the phylum Proteobacteria (e.g., species such as *Bilophila wadsworthia, Enterobacter cloacae, Enterobacter hormaechei,* and *Klebsiella aerogenes*)*. Bilophila wadsworthia, a* sulfite-reducing pathobiont, breaks down sulfur-containing organic matter and produces hydrogen sulfide (H_₂_S). Its overgrowth makes too much hydrogen sulfide, which triggers chronic inflammation, intestinal barrier breakdown, and tissue damage, making it a key pathobiont linked to various gastrointestinal and metabolic diseases. This bacterium is linked to a diet high in saturated (milk-derived) fat but not in polyunsaturated (safflower oil) fat [56,57]. *Enterobacter cloacae* complex, including *Enterobacter cloacae* and *Enterobacter hormaechei,* can proliferate abnormally in the gut of individuals with obesity, inflammatory bowel disease, or metabolic disorders, and can exacerbate metabolic disease by inducing inflammation and lipid accumulation [58]. *Enterocloster bolteae* has been reported to produce butyrate, propionate, and acetate, which alter gastrointestinal motility and contraction rates, contributing to chronic diarrheal episodes [59]. *E. bolteae* appears to produce *beta*-lactamases with resistance to β-lactam antibiotics such as penicillin G, ticarcillin, and piperacillin–tazobactam [60]. Frequent antibiotic use in urban settings can reduce microbial diversity by eliminating both harmful and beneficial bacteria [61]. Interestingly, rural residents reported higher antibiotic use in the prior 3 months than those living in urban areas in our study. However, our main findings were not significantly changed in the sensitivity analysis excluding 29 residents who reported antibiotic use in the prior 3 months (Data not shown). Although not measured in our study, air pollution should not be ignored, especially in Hanoi, which has a notable urban air quality profile [44]. Air pollution has also been linked to systemic inflammation and to alterations in gut microbial composition [62].

Most of the above-mentioned residence-related microbiome differences were attenuated after further adjustment for socioeconomic, lifestyle, and dietary factors, as well as comorbidity, suggesting that these factors shape the microbiome among urban and rural residents. Of note, there were a few significant associations that remained after the full adjustments, including the enrichment in species *Prevotella pectinovora*, strain *SGB1662*, species *Ruminococcus_sp_AF13_28,* strain *SGB1662,* class *CFGB1474*, order *OFGB1474*, and family *FGB1474,* genus *GGB3731,* species *GGB3731_SGB5063,* strain SGB*5063,* genus *GGB4456,* species *GGB4456_SGB6141* and Strain *SGB6141* class *CFGB1458,* order *OFGB1458,* family *FGB1458,* and phylum *Actinobacteria* among the rural residents (FDR <0.1). Species *Prevotella pectinovora*, an anaerobic bacterium characterized as a novel pectin-degrading species capable of utilizing pectin and other plant-derived polysaccharides, supports a role in carbohydrate degradation [63]. Additionally, the species *Ruminococcus_sp_AF13_28,* as well as most *Ruminococcus* species, are involved in the metabolism of human N- and O-glycans, and in fiber metabolism into short-chain fatty acids (SCFAs) like acetate and butyrate [64]. Our findings suggested that higher rice and fiber consumption can promote the selection of carbohydrate-degrading *Prevotella* and fiber-degrading *Ruminococcus*, potentially supporting health-related activities. Notably, genus *Adlercreutzia*, species *Adlercreutzia equolifaciens*, and strain *SGB14798* were significant only at FDR<0.1 in Model 2 when adjusted for socioeconomic characteristics, lifestyle, and dietary factors, but they lost statistical significance in the fully adjusted Model 3. Species *Adlercreutzia equolifaciens* is capable of producing equol, a metabolite widely reported to be associated with improvements in obesity and colitis. Additionally, *A. equolifaciens* may enhance a healthy gut microbiome and host health by promoting the production of beneficial metabolites, such as 3-indoleacrylic acid and palmitoylserinol [65]. More studies are needed to understand the attributes of these differences.

We found suggestive evidence of residence-related differences in gut microbiome-mediated degradation pathways for L-glutamate/propanoate and sucrose. These findings are biologically plausible in light of the dietary and activity differences observed by residence. These microbial metabolic pathway results should be viewed as candidate functional hypotheses to be prioritized for replication rather than as definitive conclusions.

This is the first study in Vietnam to evaluate differences in gut microbiome profiles between women living in urban and rural areas using shotgun metagenomic data. Gut microbial composition profiling was performed using MetaPhlAn 4.1.1, which identified from ∼1M microbial genomes (∼236,600 references and 771,500 metagenomic assembled genomes) spanning 26,970 species-level genome bins. This allowed us to estimate the prevalence and relative abundance of species or strains with enhanced resolution and accuracy [31]. In addition, we comprehensively evaluated gut microbiome profiles using multiple diversity metrics, individual microbial taxa and metabolic pathways, and staged adjustment models to distinguish age-adjusted residence patterns from associations that persist after adjustment for measured covariates. Last but not least, we classified residence based on long-term geographic exposure (i.e., prior 10-year residence) and used a detailed survey.

Several limitations should also be noted. The cross-sectional design precludes temporal or causal inference. The sample size was modest, which limits power for assessing high-dimensional microbiome features and increases the instability of effect estimates. Influences of environmental and some unmatched socioeconomic dimensions were not assessed. Dietary variables were based on frequency categories rather than detailed nutrient or food-weight data, and medication exposure did not capture timing, dose, or indication. Multiple testing is a major challenge for microbial taxa and metabolic pathway analyses, likely contributing to non-significant findings, particularly in the latter due to the limited sample size. Finally, only middle-aged female residents were included in this study. Thus, the results may not generalize to men, other age groups, or urban and rural residents with different diets, environments, or health care systems.

In conclusion, we found differences in the gut microbiome between urban and rural residents in Vietnam that may be attributable to urbanization-related changes in lifestyle, diet, and comorbidity exposures. We also identified candidate microbial and functional features for validation in further larger, longitudinal studies.

## Acknowledgments

The authors would like to give special thanks to the project participants and research staff members, without whom this study would not have been possible. This work was conducted in part using the resources of the Advanced Computing Center for Research and Education at Vanderbilt University, Nashville, TN, USA.

## Author contribution

Conceptualization, JG, XOS, and SMN; Methodology, JG, XOS, and SMN; Writing— original draft preparation, JG, HTTT, QC, MVH, TVT, XOS and SMN; Data curation, SMN, HTTT, and QC; Writing—review and editing, JG, HTTT, QC, MVH, TVT, XOS and SMN; Supervision, SMN and XOS; Project administration, HTTT, MVH, XOS and SMN; Funding acquisition, TVT, XOS and SMN. All authors have read and agreed to the published version of the manuscript.

## Data Availability Statement

Study data are available on request. The data underlying this article will be shared upon reasonable request to the corresponding author.

## Ethics Approval Statement

The study was conducted in accordance with the guidelines of the Declaration of Helsinki and approved by the Institutional Review Boards of Vietnam National Cancer Institute (No.160530; IRB approval issued May 30, 2016) and Vanderbilt University Medical Center (No.161039 “Vietnam Center of Research Excellence (V-CORE) - IRB approval issued May 31, 2018”).

## Informed Consent Statement

Written consent was obtained from all participants.

